# Historic trends and future projections of the prevalence of adult excess weight in Scotland, 2003 to 2040: a Bayesian age-period-cohort modelling study

**DOI:** 10.1101/2025.01.07.24319409

**Authors:** Robby De Pauw, Fatim Lakha, Eilidh Fletcher, Diane L Stockton, Emma Baird, Suzanne Connolly, Brecht Devleesschauwer, Grant M A Wyper

## Abstract

**Introduction:** The prevalence of excess weight in Scotland is higher than other UK nations and amongst the highest when compared with European Union countries. The aim of our study was to use historic data to identify and integrate age-period-cohort (APC) effects into projected estimates of the prevalence of excess weight in Scotland.

**Methods:** Interviewer-validated height and weight measurements were obtained for 72,542 adults between 2003 and 2019 from the Scottish Health Survey to calculate body mass index (BMI). Relevant socio-demographic data were also sourced to contribute to modelling and results stratification. Study outcomes were defined as prevalent cases of overweight (BMI ≥ 25) and obesity (BMI ≥ 30). We estimated historic trends in study outcomes by sex and age-group, and projected future trends to 2040, using Bayesian hierarchical APC modelling.

**Results:** In 2040, we estimate 3.2 million prevalent adult cases of overweight, of which 1.5 million are cases of obesity. Projections were more pronounced for obesity, compared to overweight, with around an additional 32,000 male cases and 133,000 female cases between 2019 and 2040. Between 2003 and 2019, the proportion of male and female cases of both overweight and obesity aged 65 years and above have increased, a trend projected to further intensify as we move towards 2040.

**Conclusion:** Left unaddressed, we estimate a substantial increase in adult excess weight in Scotland by 2040, particularly for females, compounded by increases in obesity and in the proportion of older cases. These findings are a warning signal of future adverse population health impacts and healthcare service sustainability pressures. Projected estimates are not inevitable and underscore the need to accelerate progress on implementing preventative measures to address the food environment, and on further development of weight management and support services, to improve the health and wellbeing of the Scottish population.

**Key messages:** *What is already known on this topic:*

- The prevalence of excess weight in Scotland is higher than other UK nations and amongst the highest when compared with European Union countries.

*What this study adds:*

- In 2040, we estimate approximately 3.2 million prevalent adult cases of overweight in Scotland, of which 1.5 million are cases of obesity.
- Our study highlights that projections were more pronounced for obesity, compared to overweight, with around 32,000 additional male cases and 133,000 additional female cases between 2019 and 2040.
- Underpinning these findings were major shifts in the age distribution of cases. Between 2003 and 2019, the proportion of male and female cases of both overweight and obesity aged 65 years and above have increased, a trend projected to further intensify as we move towards 2040.

*How this study might affect research, practice, or policy:*

- Our public health foresight study provides insights into a possible future. These findings are not inevitable and underscore the need to accelerate progress on implementing preventative measures to address the food environment, and on further development of weight management and support services, to improve the health and wellbeing of the Scottish population.

## Introduction

Recent estimates for Scotland indicate that around two-thirds of people are overweight, and approximately one-third are living with obesity [1]. The prevalence of excess weight is disproportionately higher in Scotland compared to other regions of the UK, and among the highest when compared with countries in the European Union [2,3]. The rates of adult obesity in Scotland’s most deprived areas persistently exceed those in the least deprived areas. The most widespread metric used for determining whether an individual’s weight is higher than recommended is a person’s body mass index (BMI) or, alternatively, the waist-to-height ratio [4]. Excess weight is associated with increased risk of a wide range of common health conditions, including the risk of mortality, from conditions such as: diabetes and kidney diseases; cardiovascular diseases; cancers; musculoskeletal disorders; respiratory infections and chronic diseases; digestive diseases; and neurological disorders [5]. The health impacts are substantial with approximately 10% of all-cause health loss in Scotland attributable to excess weight [6]. In addition to preventing many people in Scotland from living longer lives in better health, they present substantial demands and pressures on health and social care services. A recent estimate placed the annual cost of obesity in Scotland at £5.3 billion, with the largest share from associated reductions in quality of life from living with obesity [7]. These costs do not consider costs attributable to overweight excluding obesity, which is highly prevalent and remains associated with significant adverse health outcomes, indicating that the cost of excess weight is likely to be considerably more.

Successive Scottish Government policies including 2018’s ‘*A healthier future: Scotland’s diet and healthy weight delivery plan*’ have laid the foundations for clear areas for action to address excess weight [8]. The 2018 delivery plan aims to transform the food environment by ensuring: (i) children have the best start in life; (ii) healthier choices are supported by the food environment; (iii) people have access to effective weight management services; (iv) leaders across public and private sectors promote healthy diets and weight; and (v) diet-related health inequalities are reduced. Since the publication of this delivery plan, public services have faced increasing pressures during the COVID-19 pandemic and beyond. Findings from the Scottish Burden of Disease study illustrate that, if left unaddressed, changes in population and ageing would be expected to significantly increase disease burdens by around 21% by 2040, placing intense demands on the healthcare system [9]. This highlights the need, and importance, of utilising epidemiological methods which allow us to anticipate future trends; not only in the occurrence of health conditions, but also from associated health risk exposures such as high BMI [10]. Public health foresight methods provide us with a means to engage with, and plan for, possible futures. Scotland has a longstanding annual survey which collects validated data on the BMI of respondents, which presents a key opportunity to harness this data to undertake age-period-cohort (APC) analysis [11]. APC analysis allow us to separate, and project trends of, effects related to: age on excess weight; how excess weight develops over time; and, the difference in the risk of excess weight in successive birth cohorts. There are several advantages to this approach, the main one being that period and cohort effects serve as proxies for events such as changes in the fundamental determinants of health, and public health and medical interventions, which are often difficult to measure directly.

The aim of our study was to use historic data to identify APC effects and integrate these into the modelling of projected estimates of the adult prevalent cases of excess weight for males and females in Scotland.

## Methods

### Study population and sampling period

The study setting was Scotland, a country with a population of approximately 5.5 million residents, of which approximately 4.6 million are aged 16 years and above [12]. The sample was restricted to adults aged 16 years and above, and the sampling period for assessing historic APC effects in the prevalence of excess weight was 2003 to 2019 with a future projection period up to 2040. The projection period was set to align with other scenario work that has been carried out as part of the Scottish Burden of Disease study [9].

### Data

Our study sourced data from the UK Data Service for 13 cross-sectional waves of the Scottish Health Survey (2003, 2008 to 2019 inclusive) [13]. The Scottish Health Survey aims to provide a detailed picture of Scotland’s population health through sampling private households using stratified cluster probability sampling of the non-institutionalised residential population, which has been described in further detail elsewhere [14]. The survey has been run annually since 2008, prior to which it was conducted intermittently. We sourced data on interviewer-validated height and weight which formed the basis of our study outcome variables. The height and weight of respondents was objectively measured by survey interviewers, allowing for the calculation of BMI 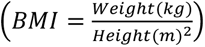. BMI was then used to define our binary study outcomes of overweight (BMI ≥ 25) and obesity (BMI ≥ 30).

The age at survey date, sex, and area-level deprivation quintile defined by the Scottish Index of Multiple Deprivation (SIMD) was sourced for survey respondents. SIMD is a small-area index of multiple deprivation and is based on approximately 7,000 data zones. It incorporates pooled and weighted indicator data from seven domains: employment; income; crime; housing; health; education; and geographical access [15]. The resulting composite SIMD ranking for each data zone is subsequently converted into quintiles, where the first quintile (SIMD 1) reflects the 20% most deprived data zones, with the fifth quintile (SIMD 5) reflecting the 20% least deprived data zones. The age at survey date was used to establish the birth year for each survey respondent. The variables entered into APC models as explanatory variables were sex, age at survey date, birth year, survey year, and SIMD quintile. Survey weights were also obtained and used to ensure that estimates were adjusted to account for variations in sampling and survey responses across each of the 13 survey waves.

Mid-year Scottish annual population estimates for 2003 to 2019 were sourced from the Office of National Statistics (ONS) by sex, single year of age, and SIMD quintile [16]. Annual population projections up to 2040 were sourced from ONS by sex and single year of age [17]. At the time of this study being undertaken, both ONS and National Records of Scotland advised use of the international migration variant 2020-based interim national population projections [18]. This was because these estimates included the most recent estimates of international migration in the averages used to calculate the short- and long-term international migration assumptions. These population estimates allow us to harness the sample estimates to generate estimates of the extent of overweight and obesity at population-level in Scotland.

### Analyses

We used hierarchical APC (HAPC) modelling to estimate the number of prevalent cases of overweight and obesity using two separate APC models. HAPC models allow for a mixture of fixed and random effects, whereas there is a linear dependency between age, period, and cohort effects in classical APC models [19]. The number of prevalent cases (*y_ij_*) for a given age-group *i* and period *j* was modelled as a Binomial process, where the mean was equal to the product of the population-at-risk (*N_ij_*) and the estimated prevalence:

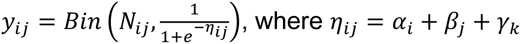

The logit of the prevalence (*η_ij_*) was estimated as the sum of the combination of age (*α_i_*), period (*β_j_*) and cohort (*γ_k_*) effects, where *k = M(l – i) + j* was the birth year cohort, *M* was the number of periods per age-group, and *l* was the number of age-groups. Further analysis was undertaken to explore the impact of the possible loss of information from modelling dichotomised outcomes by alternatively modelling the continuous variable of population-level BMI (with an identity link function) and comparing both models.

We applied second-order random walk priors (RW2) to the age, period, and cohort effects, due to their suitability when modelling unequal time intervals when estimating age, period, and cohort effects [20]. Model selections for factors and priors were based on the Deviance Information Criteria (DIC) and Watanabe–Akaike Information Criterion (WAIC). We assessed the fit and predictive accuracy of models by fitting the model to repeated cross-sectional sample surveys. The proportion of the variation in the outcome that was explained by the independent variables (R^2^) and the root mean squared error (RMSE) were estimated for different priors and models.

We describe estimates using the median, and 95% credibility intervals (CrI) around the median which were constructed based on the 2.5th and 97.5th percentile of the posterior distribution. The results presented in this paper relate to absolute numbers of prevalent cases of overweight and obesity, as our motivation for these analyses was to consider how these estimates could impact population-level needs and demands in the future. This means that our results do not control for differences in underlying population structure. However, we have also carried out analyses which compare crude and age-specific rates of excess weight, whereby confounding from different population sizes and/or structure is removed, with results consistent with our main analyses. Utilising the strengths of applying Bayesian HAPC models, we estimated the probability of scenarios of increased prevalence for overweight and obesity by 2030 and 2040, separately for males and females, where the reference was 2019 estimates [21]. The thresholds used were increases greater than: 0%, 5%, 10%, 25%, 50%, and 100%, where a 100% increase would be equivalent to double the 2019 estimate.

All statistical analyses were performed in R 4.2.0 [22]. HAPC models were fitted using the Integrated Nested Laplace Approximation (INLA) package (Version 22.05.07) [23]. The use of INLA is more efficient for Bayesian inference compared to Bayesian Markov chain Monte Carlo methods which are computer intensive.

## Results

### Study sample

The overall sample size of adults aged 16 years and above was 72,542 participants across all 13 cross-sectional waves of the Scottish Health Survey (Table S1, Supplementary Material). Generally, the sample size has decreased over time from the 2003 to the 2019 survey. The median age of participants ranged from 50 to 54 years across all survey waves, with a higher proportion of female participants. The sex distribution was consistent across survey waves, ranging from 55 to 57% female respondents. In general, the proportion of respondents from the least deprived areas slightly increased over time from 2003 to 2019, whilst the proportion of respondents from the most deprived areas slightly decreased over time, although there were some inconsistent annual fluctuations in this long-term trend.

### Contribution of factors to the prevalence of excess weight

The multivariable analysis of associated factors for excess weight, based on Bayesian APC models, for males and females are outlined in Table 1 by study outcome. The odds of overweight by sex were higher for males (Females vs Males, Odds Ratio (OR) = 0.69; 95% CrI = 0.67 to 0.72), whereas the odds of obesity were higher for females (OR = 1.07; 95% CrI = 1.03 to 1.11). We found that the odds of both overweight and obesity increased by increasing levels of area-level deprivation. These odds increased linearly for obesity from 31% higher for SIMD 4 (SIMD 4 vs SIMD 5, OR = 1.31; 95% CrI = 1.23 to 1.39) to 87% higher for the most deprived areas (SIMD 1 vs SIMD 5 OR = 1.87; 95% CrI = 1.76 to 1.98). This trend was generally similar for overweight, although at a slightly lower level, and the highest odds of overweight were estimated for those residing in SIMD 2; the second most deprived area-level deprivation quintile (SIMD 2 vs SIMD 5 OR = 1.45; 95% CrI = 1.38 to 1.53). The odds of both overweight and obesity by age followed a similar pattern and were generally highest for the ages of 35 to 64 years, and then started to lower from the ages of 65 years and above but remained higher than the under 25 years reference group until the ages of 85 years and above. We also found that the odds of overweight and obesity reduced for successive birth cohorts, with the lowest odds estimated for more recent birth cohorts. On the other hand, the odds of overweight and obesity increased by each period with the highest odds being estimated for the most recent periods (2019 vs 2003, Overweight OR = 1.54; 95% CrI = 1.52 to 1.57; Obesity OR = 1.59; 95% CrI = 1.56 to 1.62). A further visualisation of the estimated fixed and random effects for male and female models for overweight and obesity are presented in the Supplementary Material section (Figures S1 to Figure S5).

**Table 1:**
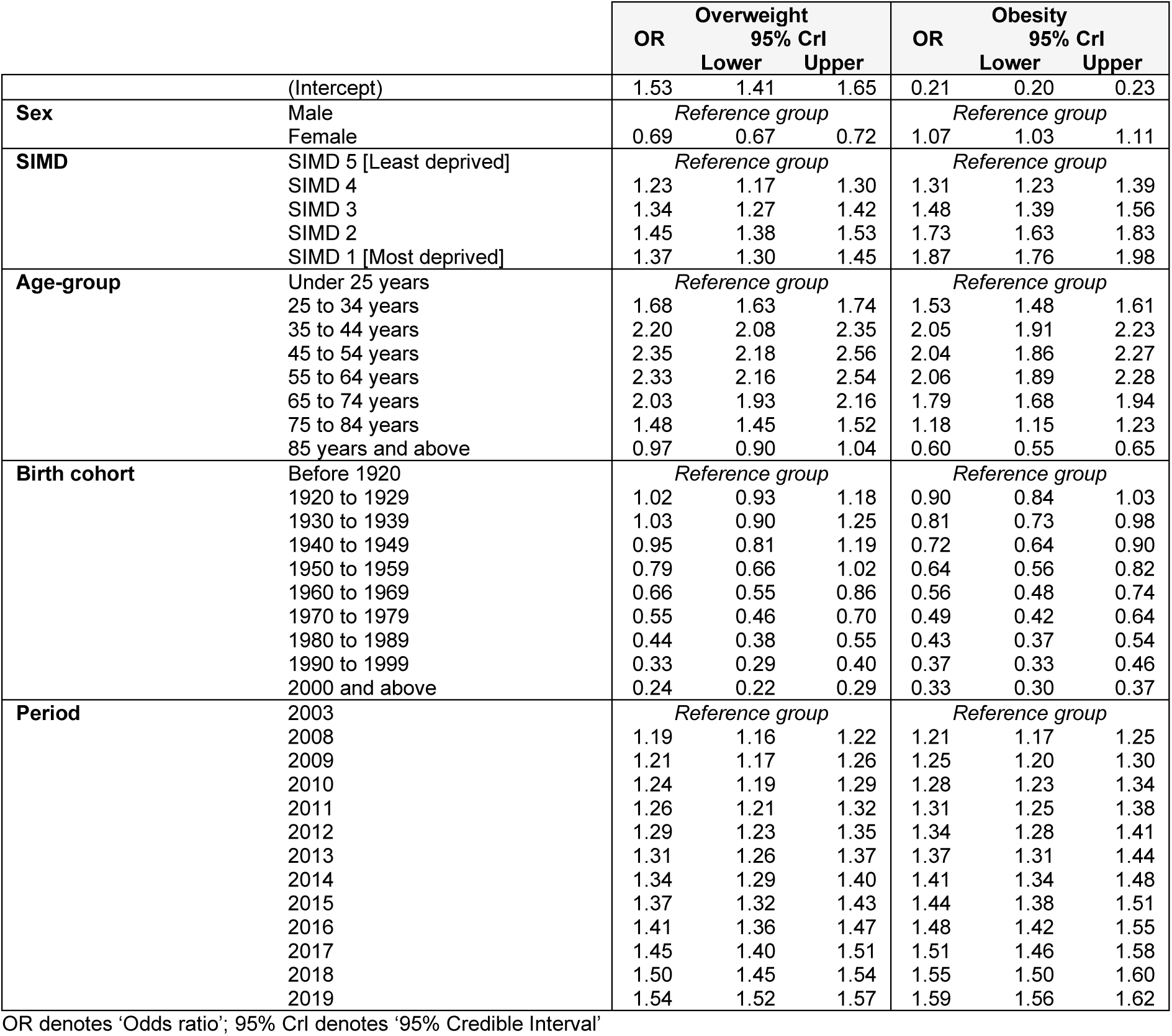
Odds ratios (OR) for excess weight in Scotland based on Bayesian age-period-cohort model of sample data by study outcome.

### Trends in the prevalence of excess weight

The estimated number of prevalent cases of overweight and obesity for males increased by 14% and 37% respectively from 2003 to 2019, with the fastest rate of increase occurring between 2003 and 2010 (Figure 1). For females, we found that the estimated number of prevalent cases of overweight and obesity increased by 13% and 25% respectively from 2003 to 2019, with the rate of increase consistent across the full study sampling period.

**Figure 1:**
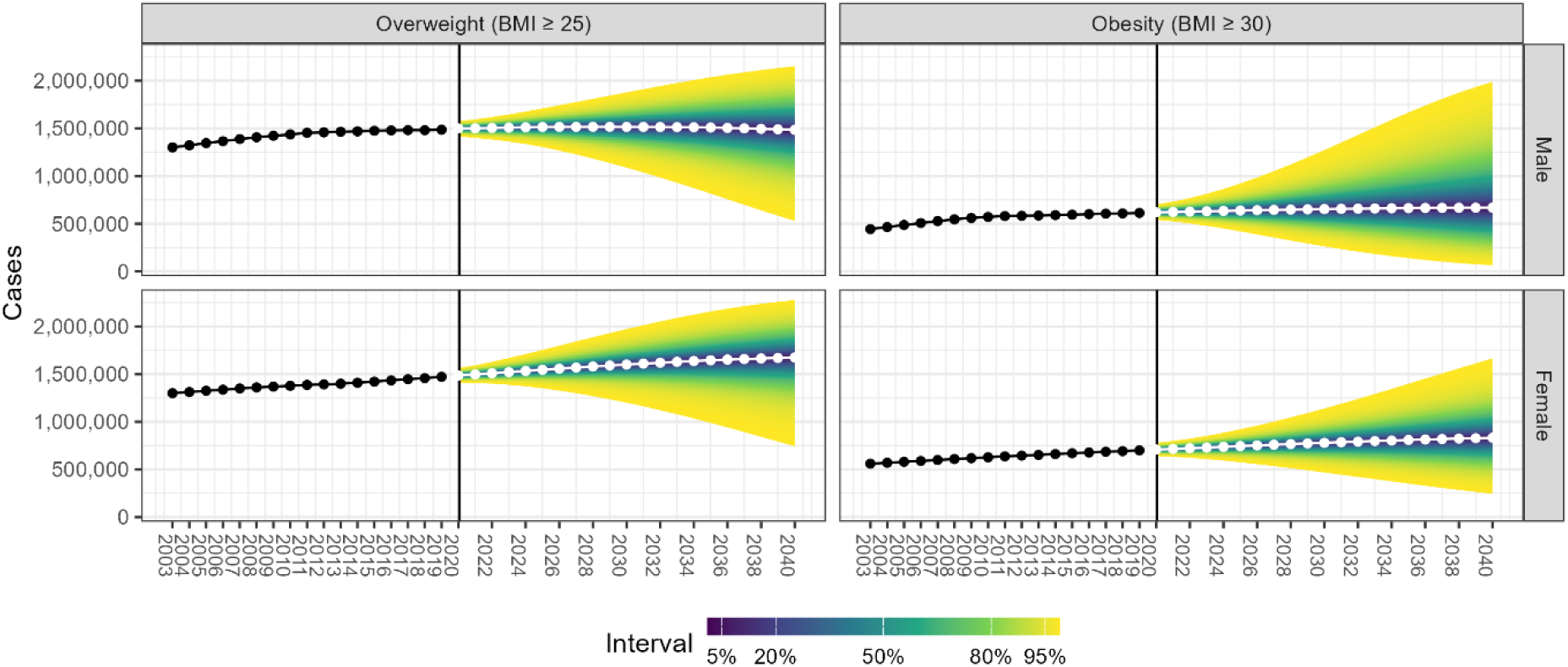
Historic and projected estimates of the number of prevalent cases of excess weight in Scotland, by sex and study outcome, 2003 to 2040

Projections indicate that the number of prevalent cases of overweight for males are estimated to increase by 2% from 2019 to 2030; an increase of approximately 29,000 cases. However, by 2040 the projected number of prevalent cases of overweight for males is estimated to return to a similar level as 2019. We estimate a larger increase of 5% in the projected number of prevalent cases of male obesity from 2019 to 2030 with cases estimated to remain at this level up to 2040, representing an increase of approximately 32,000 cases. In 2040, for males we estimate 1,482,005 (95% CrI: 814,453 to 2,004,583) prevalent cases of overweight and 642,557 (95% CrI: 140,642 to 1,624,389) prevalent cases of obesity.

The projected number of prevalent cases of overweight for females is estimated to increase by 14% from 2019 to 2040; an increase of approximately 204,000 cases, with slightly over half of this increase (63%) estimated to have occurred by 2030. We estimate larger increases of 19% in the number of female prevalent cases of obesity from 2019 to 2040; representing an increase of approximately 133,000 cases, of which around two-thirds are estimated to occur by 2030. In 2040, for females we estimate 1,674,290 (95% CrI: 1,073,990 to 2,146,288) prevalent cases of overweight and 832,480 (95% CrI: 381,915 to 1,396,634) prevalent cases of obesity.

When we assess historic sex-specific differences in the estimated number of prevalent cases of overweight, there were slightly fewer cases for females than males between 2003 and 2019. However, by 2040 we estimate 13% more prevalent cases of overweight for females compared to males. For obesity, the estimated number of prevalent cases was higher for females than males between 2003 and 2019, although the scale of difference fluctuates across the study period. We estimate 30% more prevalent cases of obesity for females compared with males by 2040.

We estimated the probability of scenarios of increased adult excess weight prevalence by 2030 and 2040, separately, for males and females (Figure 2). For males, there was an estimated 34% probability of any increase in the prevalence of overweight by 2030 and for 2040, compared to the 2019 estimate. Respective probabilities for obesity were considerably higher (2030: 50% and 2040: 49%). The estimated probabilities for females were much higher, compared with males. For females, we estimated there was a 72% probability of an increase in the prevalence of overweight by 2030 and 71% by 2040, with slightly lower respective probabilities for obesity (2030: 68% and 2040: 67%).

**Figure 2:**
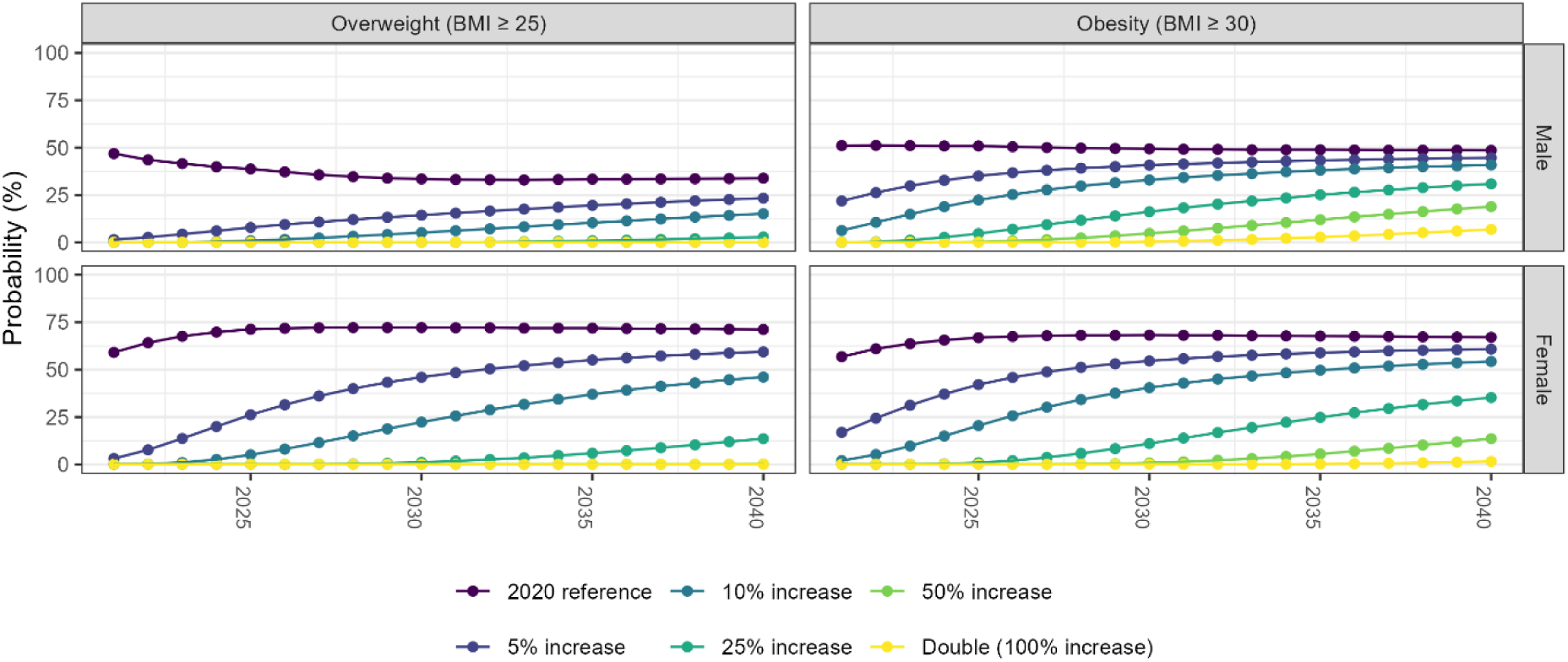
Probability of an increase in the adult prevalence of excess weight, by sex and study outcome, based on the marginal posterior distribution of future projections from the Bayesian APC analyses

The historic age structure of prevalent cases of overweight and obesity for males and females has changed from 2003 and 2019, a change which is projected to intensify further by 2040 (Figure 3). The impact of overweight and obesity in males and females aged 65 years and above has become disproportionately higher over time, relative to younger age-groups. Most prevalent cases of overweight and obesity are aged between 35 to 64 years, although there are estimated to be relatively fewer cases in this age-group by 2040 compared to the early study period due to the increasing numbers of prevalent cases aged 65 years and above.

**Figure 3:**
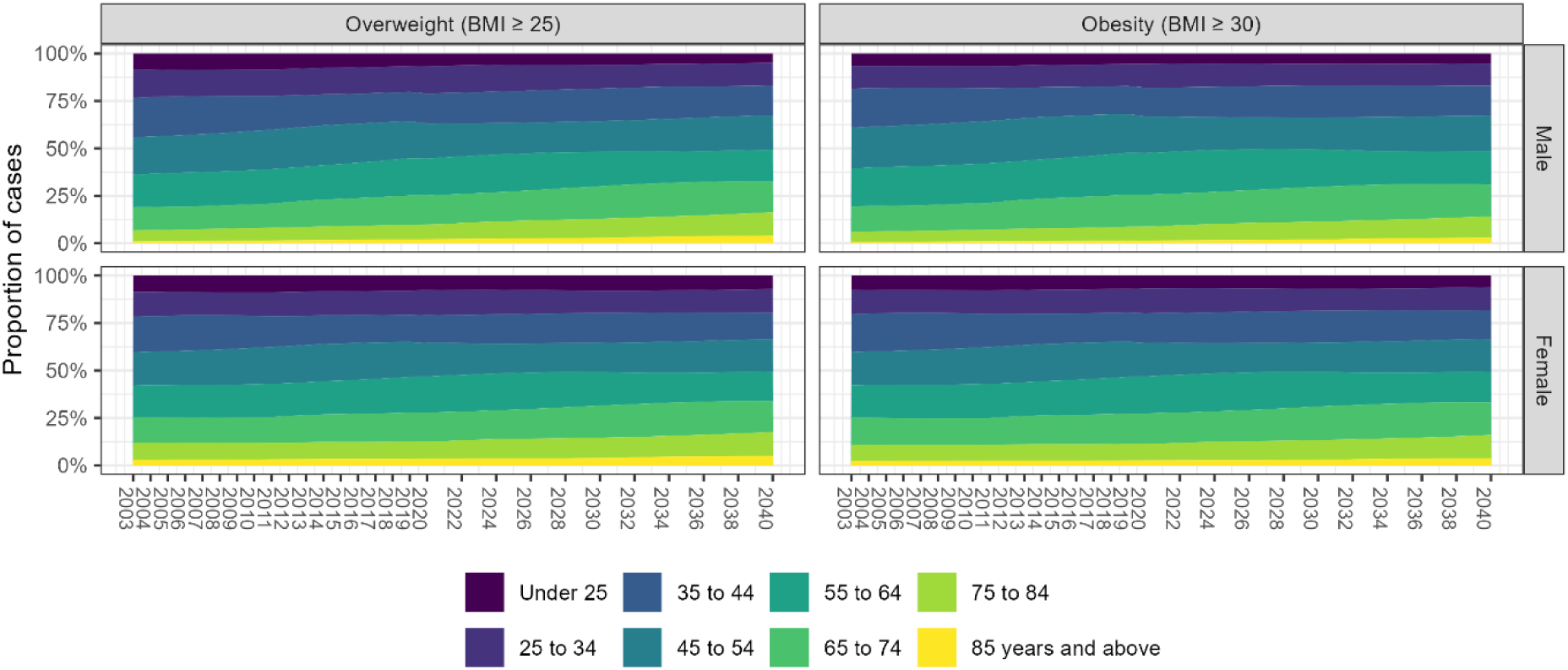
The age-relative distribution of historic and projected estimates of the number of prevalent cases of excess weight in Scotland, by sex and study outcome, 2003 to 2040

### Model validation and sensitivity analyses

When assessing the model fit, using DIC and WAIC, we found that the lowest values and therefore the most appropriate models were those that included age, period, and cohort effects, as well as the fixed effects of sex and area-level deprivation. This finding was consistent for overweight and obesity models for both males and females (Table S2, Supplementary Material). The explained variability by the overweight model was high for both males (R^2^ = 84%) and females (R^2^ = 80%). For the obesity models, the values were slightly lower (Males: R^2^ = 77%; Females: R^2^ = 67%). When we undertook a sensitivity analysis where we used BMI as the outcome variable, rather than dichotomised outcomes of overweight and obesity, the findings were compatible with that of our main analysis (Figure S6, Supplementary Material).

## Discussion

### Summary of findings

Our study estimates that in 2040 there will be approximately 3.2 million prevalent cases of overweight in Scottish adults, of which 1.5 million are cases of obesity. The number of prevalent cases of overweight is estimated to continue to increase for males until 2030 and then decrease back towards pre-pandemic levels. For females, around two-thirds of the increase in overweight cases is estimated to have occurred by 2030, a trend projected to continue to increase at a reduced rate towards 2040. Projected estimates were more pronounced for obesity, relative to overweight, with around 32,000 additional male cases and 133,000 additional female cases estimated between 2019 and 2040. A factor contributing towards these trends is the estimated impact of period effects, which are indicative of changes that affect people of all ages at a specific point in time. We estimate that with each consecutive year up 2019 the period effect becomes stronger, adversely influencing an increased number of projected cases in the future.

Historic pre-pandemic estimates illustrate there were slightly fewer prevalent cases of overweight for females compared to males, however projections in 2040 estimate 13% more female cases compared to males. There have historically been a higher number of prevalent cases of obesity for females, and by 2040 we estimate 30% more female cases compared to males. Underpinning these estimates are major shifts in the age distribution of prevalent cases. Historic estimates between 2003 and 2019 show that the proportion of male and female cases of excess weight aged 65 years and above have increased, a trend projected to further intensify as we move towards 2040.

Our methodological approach allows us to manage expectations for policymakers by communicating the probability of scenario-based increases in the prevalence of overweight and obesity. We found that by 2040, the probability of an increase in the number of prevalent cases of overweight for females was double that of males (71% vs 34%). The probability of an increase in the number of prevalent cases of obesity by 2040 was slightly lower than that of overweight but remained higher for females compared to males (67% vs 49%).

### Implications for policy and future research

The significant extent of adverse population-level health outcomes attributable to overweight and obesity are clearly evidenced, which underscores the importance of the context regarding our study findings [5, 24]. A study in Belgium, which utilised the same design as our study, found similar patterns whereby female cases were on course to have the largest increases in excess weight [10]. Projections of the prevalence of cardiometabolic health conditions to 2040 in Scotland are also consistent with our results, driven by substantial increases in cases of diabetes which the Global Burden of Disease study highlight as the outcome with the greatest disease burden attributable to excess weight [5,25]. Furthermore, most studies evidence that the increases in obesity are disproportionately higher than those of overweight which is consistent with our findings [26, 27]. We are clear on the current scale of challenge that excess weight presents [28]. Our findings outline that we are not on the right track; the number of cases of excess weight are projected to increase until 2040, with a greater number of cases of obesity which increases the likelihood of associated health harms. Exacerbating this is the shift in the age distribution of cases towards that of older ages, and cases of excess weight developing earlier in the life-course which give rise to an earlier onset of morbidity. Taken together these findings present a significant challenge because older ages are when health conditions commonly manifest at the greatest rate, with multimorbidity becoming increasingly more common, which diminish people’s quality of life and impairs their ability to live longer lives in good health [29,30]. Furthermore, there are major implications for the sustainability of publicly funded health and social care services as the projected increase in cases, and shift to an older age-distribution, will intensify demand and need for services and treatments to mitigate health conditions manifesting because of excess weight [31]. Given the impact of excess weight on the occurrence of a wide range of non-communicable diseases (NCDs), reducing the number of cases would be an effective route to progress towards achieving Sustainable Development Goal (SDG) 3.4 of reducing premature deaths from NCDs by one-third by 2030 [32]. Our estimates indicate that any prospect of making progress towards achieving SDG 3.4 by 2030 is likely to be hindered, rather than supported, by trends in the prevalence of excess weight [33].

In July 2024, Public Health Scotland and the Scottish Directors of Public Health delivered a consensus statement defining key actions to improve diet and weight in Scotland, to generate improvements in population health and wellbeing [34]. Our findings suggest an increasing period effect which shows no evidence of waning, emphasise the substantial role which commercial, economic, and environment factors have in not only influencing current rates, but in intensifying future trends [35]. A multi-faceted approach which addresses the food environment through regulation, taxation, product reformulation, and affordability is fundamental to making progress in addressing rates of excess weight and reversing the increasing influence of period effects [36, 37]. In addition, availability of weight management and support services, providing evidence-based treatment and support for people living with obesity, will also be crucial [34]. Evidence indicates that around half of the Scottish public would support the taxation, and regulation, of unhealthy food products to tackle excess weight [38].

Our study focused on projecting the prevalence of excess weight defined via BMI in Scottish adults, so did not consider those aged under 16 years. There are greater uncertainties associated with BMI for children than for adults, as measurements can be influenced by factors such as changes in growth patterns between boys and girls, particularly during puberty when growth is faster [39]. There is also greater contention over which cut-offs to use to define overweight and obesity, compared with adults [40]. The best available estimates for Scotland indicate that there are substantial concerns regarding excess weight in children, with around one-third of those starting primary school being at risk of overweight and those in Scotland’s most deprived communities twice as likely than those in the least deprived communities; a gap which has widened in recent years [34]. Given the increasing adverse influence of period effects we estimated, we would hypothesise these would also be a key consideration for those aged under 16 years. Further research is warranted to establish whether period effects are having an increasing influence in the household and family settings, or whether the chain is being broken and increasingly children and young people are being enabled to get the best start in life, independent of the weight status of other members of their residency.

### Strengths and limitations

Our study used the best available data on BMI in Scotland from 2003 to 2019, which was based on interviewer-validated height and weight of survey respondents. This means that recorded data which were used in this study are free of any biases related to individual’s being less willing to report information due to the societal stigma related to excess weight [41]. This is a major strength given that previous studies show that people tend to report lower BMI values when self-reporting, including a study in Belgium which reported an absolute difference of 0.97 kg/m^2^ between objectively measured BMI and BMI related to self-reported measurements [41,42]. The repeated cross-sectional nature of our study sample allowed us to undertake, and establish, age, period, and cohort effects related to the prevalence of excess weight, despite having unequal periods between the first survey used in 2003 and the next in 2008 [19].

Despite being the best available source, there are some important limitations to outline. More recent Scottish Health Surveys allow for the sampling of multiple adults per household which removes the need for selection weights, however the sampling design may lead to patterns of weight clustering within households. The survey response rate of adults has decreased over time, which may introduce systematic biases if the prevalence of excess weight is different in non-respondents compared to respondents [43,44,45]. To address the impact of this limitation, we utilised survey weights to correct for differences in sampling and response rates. Whilst we do not have evidence directly related to excess weight, there is evidence that alcohol-related health harms in non-respondents are higher than those of respondents [46]. Given that inequalities in overweight and obesity are less pronounced than those which are alcohol-related, we would hypothesise that any bias is more likely to be lower for excess weight than have previously been found for alcohol consumption and harms. The dichotomisation of outcomes based on continuous variables can result in the loss of information [47]. To address this problem, we undertook a sensitivity analysis using BMI as the study outcome and observed male and female trends consistent with our main findings, indicating that the dichotomisation of outcomes did not influence our study findings. Our study sample was based on data up to 2019 since this was the most up-to-date data available in the UK Data Service at time of carrying out the analyses.

There are several advantages to the APC study design we used, the main one being that period and cohort effects serve as proxies for events such as risk factors, public health and medical interventions, which are often difficult to measure directly [48]. Projections inherently include an increased level of uncertainty as they will be influenced by changes we have not yet experienced, so thus remain susceptible to Hume’s problem of induction as these yet to be experienced events could influence the over- or under-estimation of the actual number of prevalent cases [49]. The key assumption underlying these estimates is that APC trends in the future will continue based on observed historical trends, and the validity of this assumption cannot be tested until we are able to collect these data in the future. Nevertheless, public health foresight analysis and associated uncertainty is an important tool for effective planning of policy and medical interventions development and implementation [11]. Our study utilised HAPC models which allow for a mixture of fixed and random effects, whereas there is a linear dependency between age, period, and cohort effects in classical APC models. Furthermore, the application of Bayesian HAPC models permit the direct interpretation of future estimates in credibility terms, for example we can estimate how likely it is that the prevalence of overweight and obesity will increase by at least 10% [18]. This is a strength over applying APC models in a frequentist sense, as frequentist confidence intervals cannot be interpreted in terms of credibility. This strengthens the potential of these study findings in managing expectations when being used to inform discussions over improving these trends.

We found that successive birth cohorts had a reduced risk of overweight and obesity, when isolated from other effects. One such explanation supporting these results could be related to successive birth cohorts being increasingly exposed to the risks posed from excess weight on health, and increased education and awareness over how to reduce the risk of associated harms. Nevertheless, it remains tricky to interpret the role of birth cohort effects without considering how period, and age, effects are influencing rates of excess weight, as well as the fixed effects. The estimated adversely increasing period effects are an important warning as they indicate that tackling the impact of commercial, economic, and environmental determinants of excess weight are likely to be the most effective means of improving prevalence rates. Since those who have been overweight and living with obesity for long periods of time are at greater risk of premature mortality than those who are not, we hypothesise that this would at least partially explain why we found that age effects reduce for age-groups 65 years and above.

## Conclusion

Left unaddressed, we estimate a substantial increase in adult excess weight in Scotland by 2040, particularly for females, compounded by increases in obesity and in the proportion of older cases. These estimates are not inevitable and underscore the need to accelerate progress towards a multi-faceted prevention-focused approach. Further development of weight management and support services, providing evidence-based treatment and support for people living with obesity, will be important. However, crucially, the key focus must be on improving the food environment through regulation, taxation, product reformulation, and affordability.

## Article information

## Abbreviations

APC: Age-period-cohort
BMI: Body mass index
COVID-19: Coronavirus Disease 2019 CrI Credibility Intervals
DIC: Deviance Information Criteria
HAPC: Hierarchical age-period-cohort
INLA: Integrated Nested Laplace Approximation kg Kilogram
M: Metre
NRS: National Records of Scotland
ONS: Office for National Statistics
OR: Odds ratio
RW2: Second-order random walk priors
SIMD: Scottish Index of Multiple Deprivation
UK: United Kingdom
WAIC: Watanabe–Akaike Information Criterion

## Acknowledgements

We are grateful to the interviewers and participants involved in the Scottish Health Survey, and the inclusion of this data in the UK Data Service. We honour the memory of Dr Ian Grant, a distinguished Scottish public health researcher, who contributed towards the generation of the idea and design of this study, but sadly passed away during its implementation.

## Contributions

GMAW, RDP, BD, FL and EF generated the idea for the study. GMAW, RDP and BD developed the methodological approach. RDP carried out all analyses and visualisation of results. GMAW coordinated the study and drafted the original manuscript. All other authors provided critical input into the interpretation of the results, revisions to the manuscript, and approved the final draft.

## Data availability

The data that support the findings of this study can be accessed via the UK Data Service – https://ukdataservice.ac.uk/. The UK Data Service provides trusted access and training to use the UK’s largest collection of economic, population and social research data for teaching, learning and public benefit.

## Competing interests

The authors declare no competing interests.

## Funding

This research was funded by Public Health Scotland.

## Supplementary Material

**Table S1:**
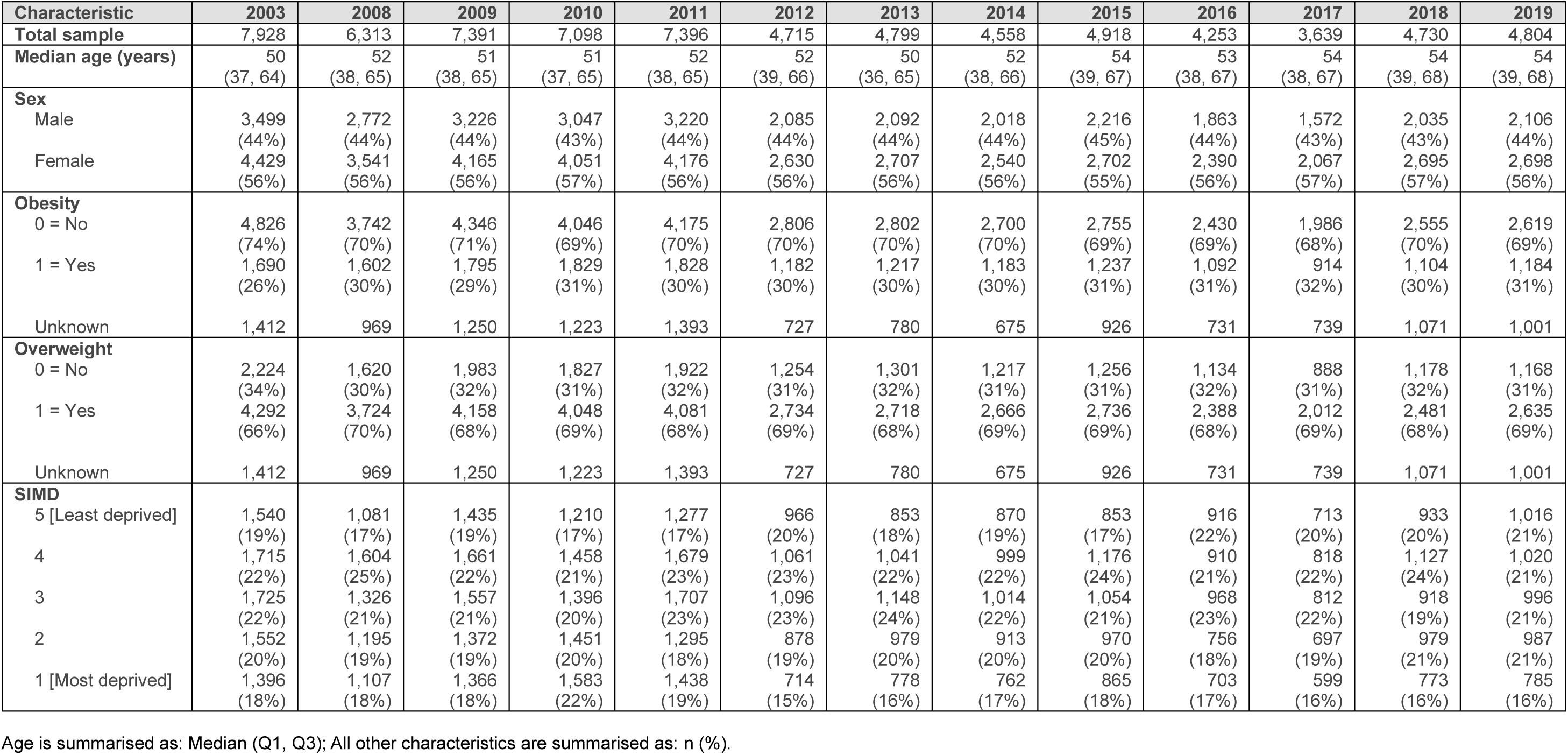
Characteristics of the study sample, Scotland, 2003 to 2019.

| Characteristic | 2003 | 2008 | 2009 | 2010 | 2011 | 2012 | 2013 | 2014 | 2015 | 2016 | 2017 | 2018 | 2019 |
| --- | --- | --- | --- | --- | --- | --- | --- | --- | --- | --- | --- | --- | --- |
| **Total sample** | 7,928 | 6,313 | 7,391 | 7,098 | 7,396 | 4,715 | 4,799 | 4,558 | 4,918 | 4,253 | 3,639 | 4,730 | 4,804 |
| **Median age (years)** | 50<br>(37, 64) | 52<br>(38, 65) | 51<br>(38, 65) | 51<br>(37, 65) | 52<br>(38, 65) | 52<br>(39, 66) | 50<br>(36, 65) | 52<br>(38, 66) | 54<br>(39, 67) | 53<br>(38, 67) | 54<br>(38, 67) | 54<br>(39, 68) | 54<br>(39, 68) |
| **Sex** |  |  |  |  |  |  |  |  |  |  |  |  |  |
| Male | 3,499<br>(44%) | 2,772<br>(44%) | 3,226<br>(44%) | 3,047<br>(43%) | 3,220<br>(44%) | 2,085<br>(44%) | 2,092<br>(44%) | 2,018<br>(44%) | 2,216<br>(45%) | 1,863<br>(44%) | 1,572<br>(43%) | 2,035<br>(43%) | 2,106<br>(44%) |
| Female | 4,429<br>(56%) | 3,541<br>(56%) | 4,165<br>(56%) | 4,051<br>(57%) | 4,176<br>(56%) | 2,630<br>(56%) | 2,707<br>(56%) | 2,540<br>(56%) | 2,702<br>(55%) | 2,390<br>(56%) | 2,067<br>(57%) | 2,695<br>(57%) | 2,698<br>(56%) |
| **Obesity** |  |  |  |  |  |  |  |  |  |  |  |  |  |
| 0 = No | 4,826<br>(74%) | 3,742<br>(70%) | 4,346<br>(71%) | 4,046<br>(69%) | 4,175<br>(70%) | 2,806<br>(70%) | 2,802<br>(70%) | 2,700<br>(70%) | 2,755<br>(69%) | 2,430<br>(69%) | 1,986<br>(68%) | 2,555<br>(70%) | 2,619<br>(69%) |
| 1 = Yes | 1,690<br>(26%) | 1,602<br>(30%) | 1,795<br>(29%) | 1,829<br>(31%) | 1,828<br>(30%) | 1,182<br>(30%) | 1,217<br>(30%) | 1,183<br>(30%) | 1,237<br>(31%) | 1,092<br>(31%) | 914<br>(32%) | 1,104<br>(30%) | 1,184<br>(31%) |
| Unknown | 1,412 | 969 | 1,250 | 1,223 | 1,393 | 727 | 780 | 675 | 926 | 731 | 739 | 1,071 | 1,001 |
| **Overweight** |  |  |  |  |  |  |  |  |  |  |  |  |  |
| 0 = No | 2,224<br>(34%) | 1,620<br>(30%) | 1,983<br>(32%) | 1,827<br>(31%) | 1,922<br>(32%) | 1,254<br>(31%) | 1,301<br>(32%) | 1,217<br>(31%) | 1,256<br>(31%) | 1,134<br>(32%) | 888<br>(31%) | 1,178<br>(32%) | 1,168<br>(31%) |
| 1 = Yes | 4,292<br>(66%) | 3,724<br>(70%) | 4,158<br>(68%) | 4,048<br>(69%) | 4,081<br>(68%) | 2,734<br>(69%) | 2,718<br>(68%) | 2,666<br>(69%) | 2,736<br>(69%) | 2,388<br>(68%) | 2,012<br>(69%) | 2,481<br>(68%) | 2,635<br>(69%) |
| Unknown | 1,412 | 969 | 1,250 | 1,223 | 1,393 | 727 | 780 | 675 | 926 | 731 | 739 | 1,071 | 1,001 |
| **SIMD** |  |  |  |  |  |  |  |  |  |  |  |  |  |
| 5 [Least deprived] | 1,540<br>(19%) | 1,081<br>(17%) | 1,435<br>(19%) | 1,210<br>(17%) | 1,277<br>(17%) | 966<br>(20%) | 853<br>(18%) | 870<br>(19%) | 853<br>(17%) | 916<br>(22%) | 713<br>(20%) | 933<br>(20%) | 1,016<br>(21%) |
| 4 | 1,715<br>(22%) | 1,604<br>(25%) | 1,661<br>(22%) | 1,458<br>(21%) | 1,679<br>(23%) | 1,061<br>(23%) | 1,041<br>(22%) | 999<br>(22%) | 1,176<br>(24%) | 910<br>(21%) | 818<br>(22%) | 1,127<br>(24%) | 1,020<br>(21%) |
| 3 | 1,725<br>(22%) | 1,326<br>(21%) | 1,557<br>(21%) | 1,396<br>(20%) | 1,707<br>(23%) | 1,096<br>(23%) | 1,148<br>(24%) | 1,014<br>(22%) | 1,054<br>(21%) | 968<br>(23%) | 812<br>(22%) | 918<br>(19%) | 996<br>(21%) |
| 2 | 1,552<br>(20%) | 1,195<br>(19%) | 1,372<br>(19%) | 1,451<br>(20%) | 1,295<br>(18%) | 878<br>(19%) | 979<br>(20%) | 913<br>(20%) | 970<br>(20%) | 756<br>(18%) | 697<br>(19%) | 979<br>(21%) | 987<br>(21%) |
| 1 [Most deprived] | 1,396<br>(18%) | 1,107<br>(18%) | 1,366<br>(18%) | 1,583<br>(22%) | 1,438<br>(19%) | 714<br>(15%) | 778<br>(16%) | 762<br>(17%) | 865<br>(18%) | 703<br>(17%) | 599<br>(16%) | 773<br>(16%) | 785<br>(16%) |

Age is summarised as: Median (Q1, Q3); All other characteristics are summarised as: n (%).

**Table S2.**
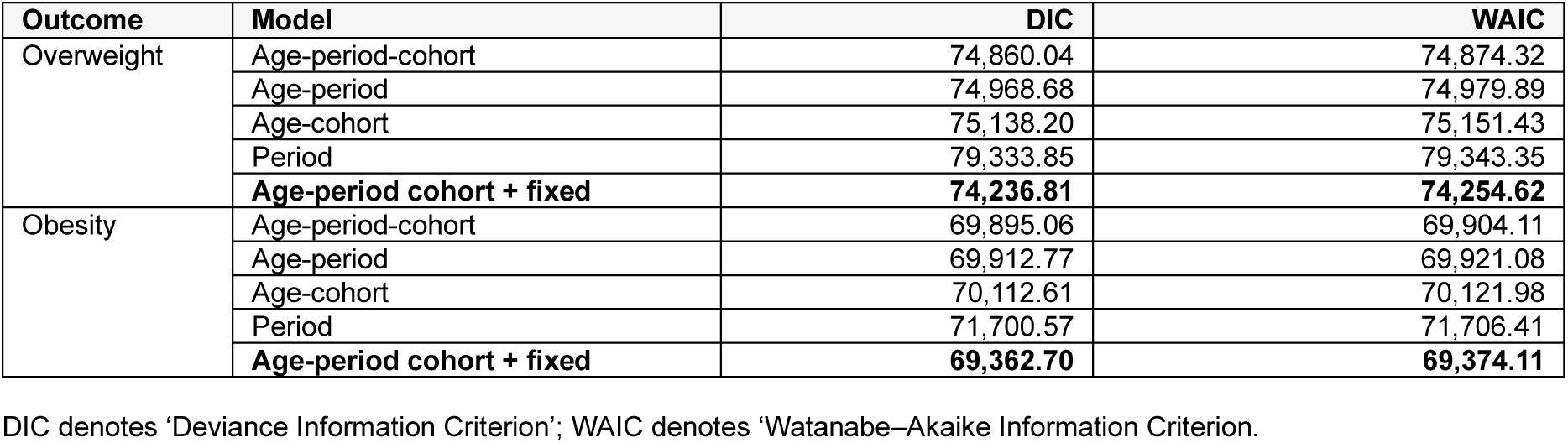
Model selection criteria for overweight and obesity, by sex.

| Outcome | Model | DIC | WAIC |
| --- | --- | --- | --- |
| Overweight | Age-period-cohort | 74,860.04 | 74,874.32 |
|  | Age-period | 74,968.68 | 74,979.89 |
|  | Age-cohort | 75,138.20 | 75,151.43 |
|  | Period | 79,333.85 | 79,343.35 |
|  | **Age-period cohort + fixed** | **74,236.81** | **74,254.62** |
| Obesity | Age-period-cohort | 69,895.06 | 69,904.11 |
|  | Age-period | 69,912.77 | 69,921.08 |
|  | Age-cohort | 70,112.61 | 70,121.98 |
|  | Period | 71,700.57 | 71,706.41 |
|  | **Age-period cohort + fixed** | **69,362.70** | **69,374.11** |

DIC denotes 'Deviance Information Criterion'; WAIC denotes 'Watanabe–Akaike Information Criterion'.

**Figure S1:**
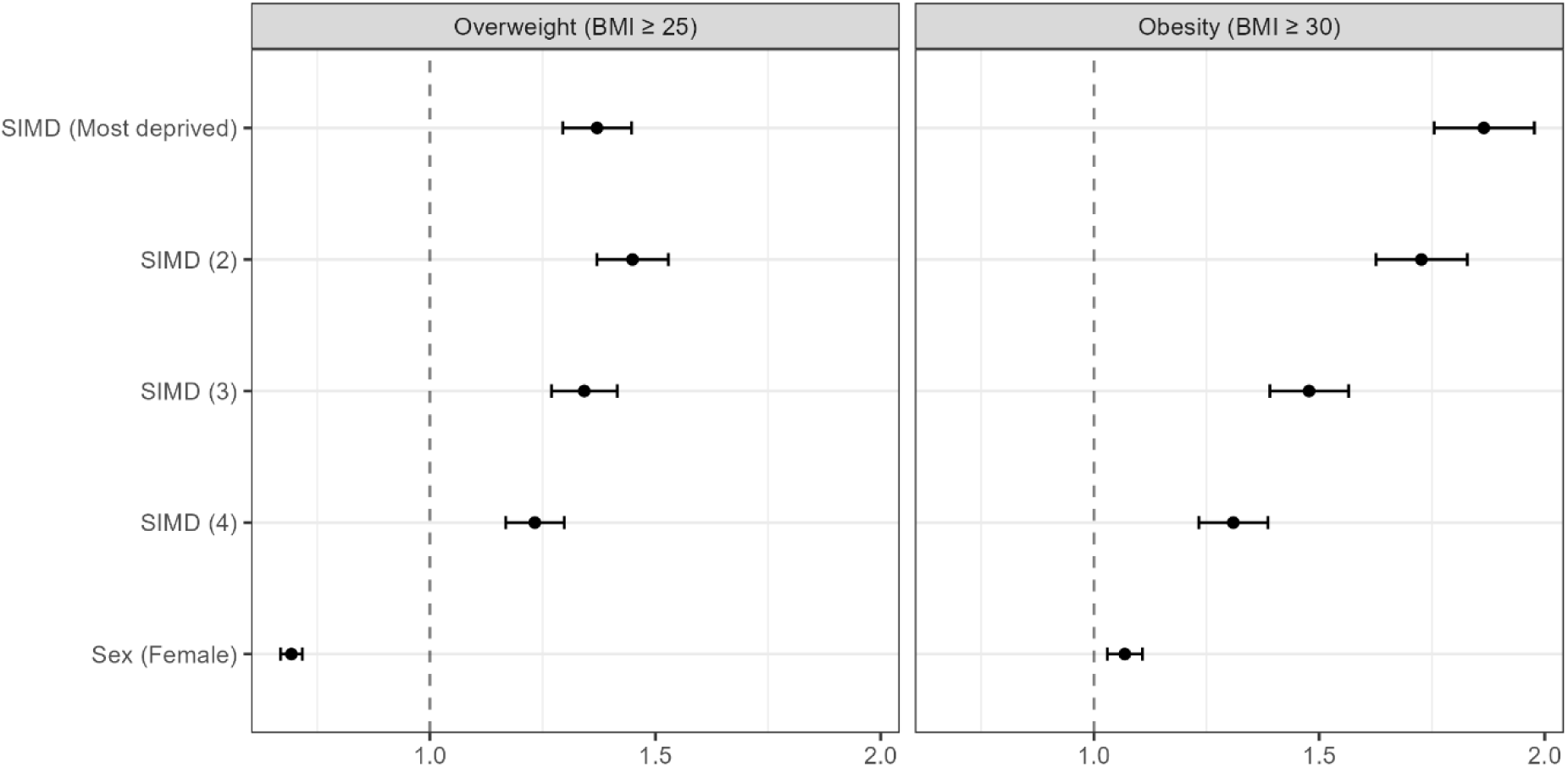
Overview of fixed effects included in the APC model by study outcome.

**Figure S2:**
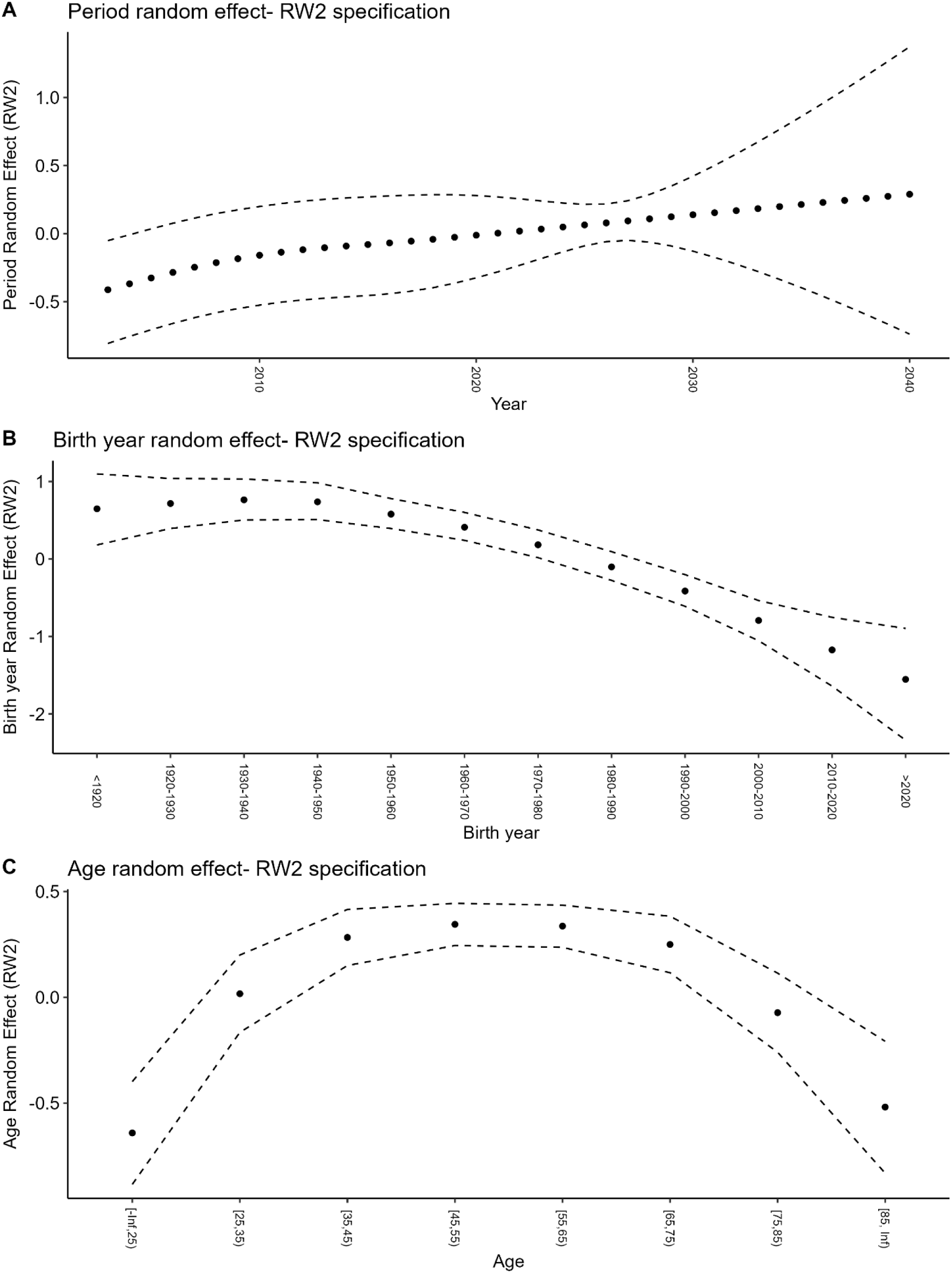
Overview of random smoothed (RW2) effects included in the male APC model for overweight.

**Figure S3:**
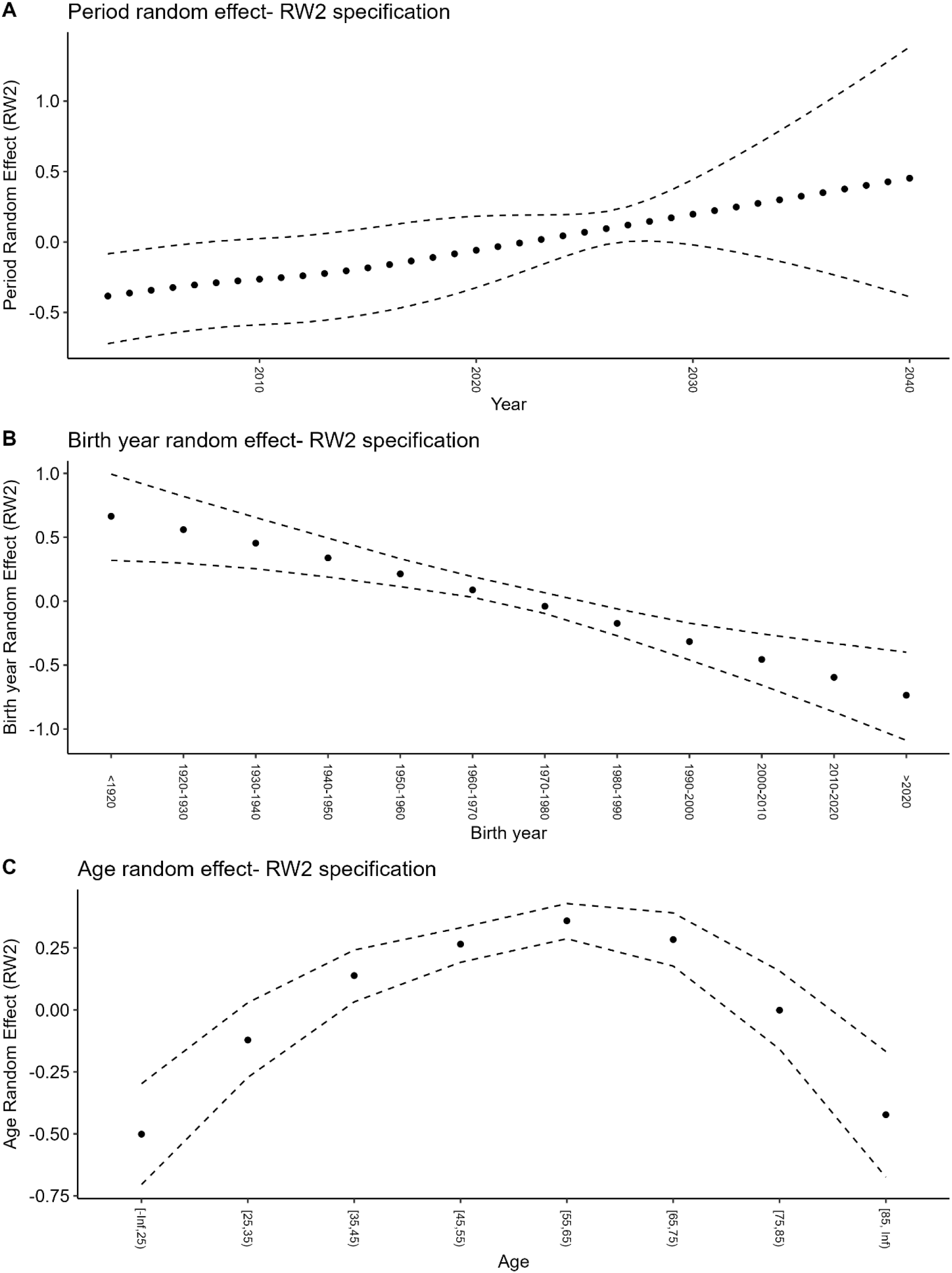
Overview of random smoothed (RW2) effects included in the female APC model for overweight.

**Figure S4:**
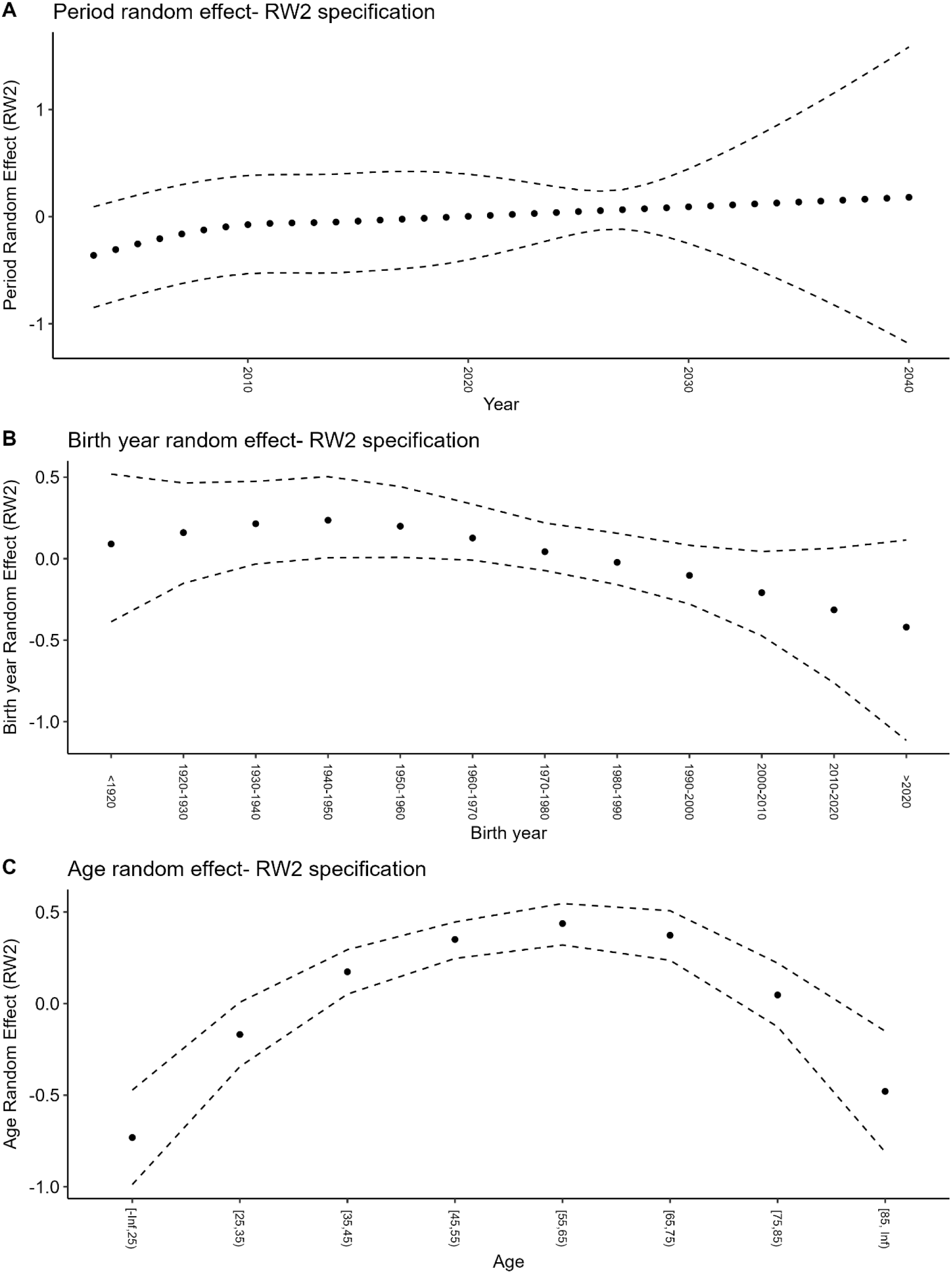
Overview of random smoothed (RW2) effects included in the male APC model for obesity.

**Figure S5:**
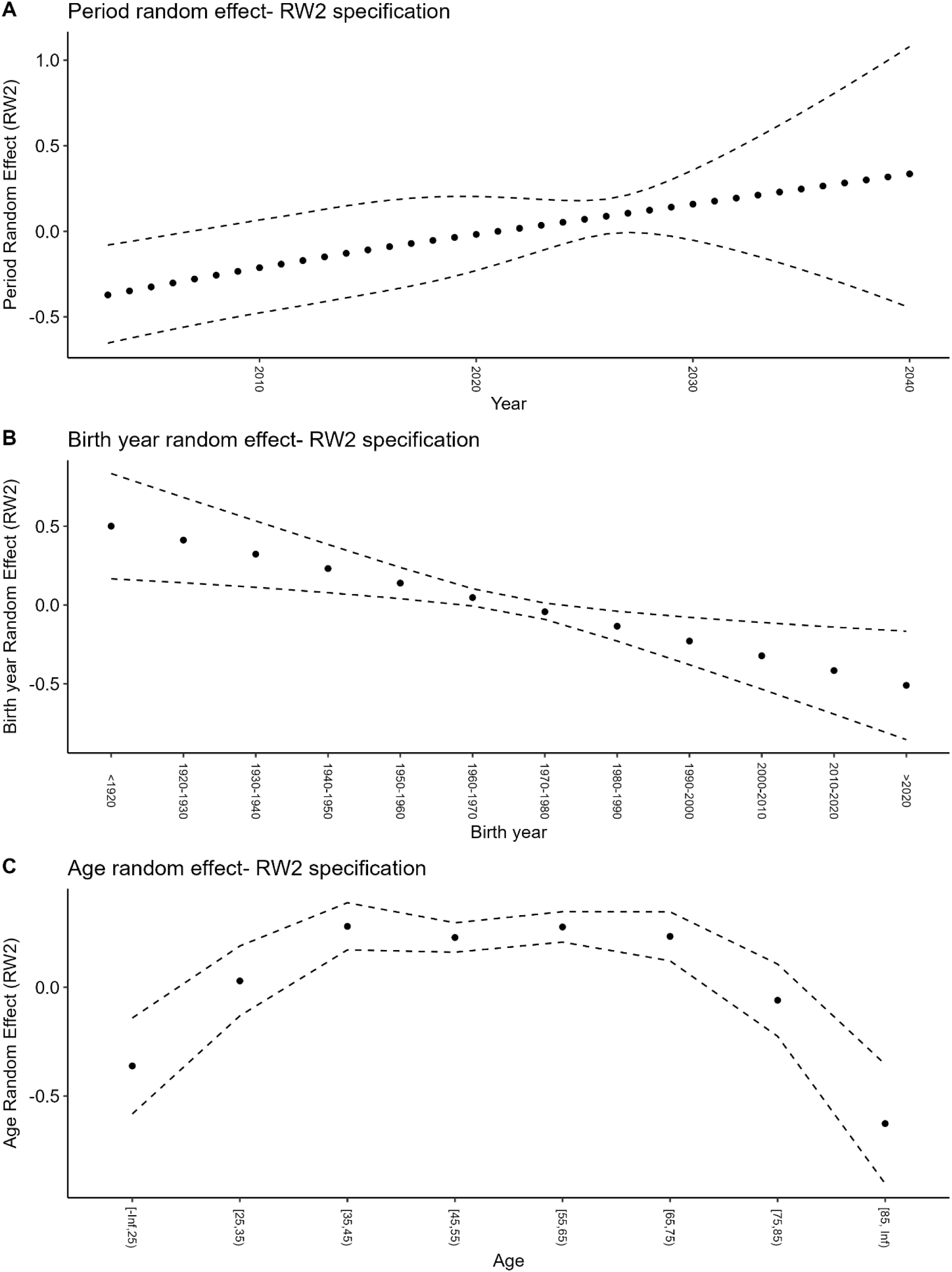
Overview of random smoothed (RW2) effects included in the female APC model for obesity.

**Figure S6:**
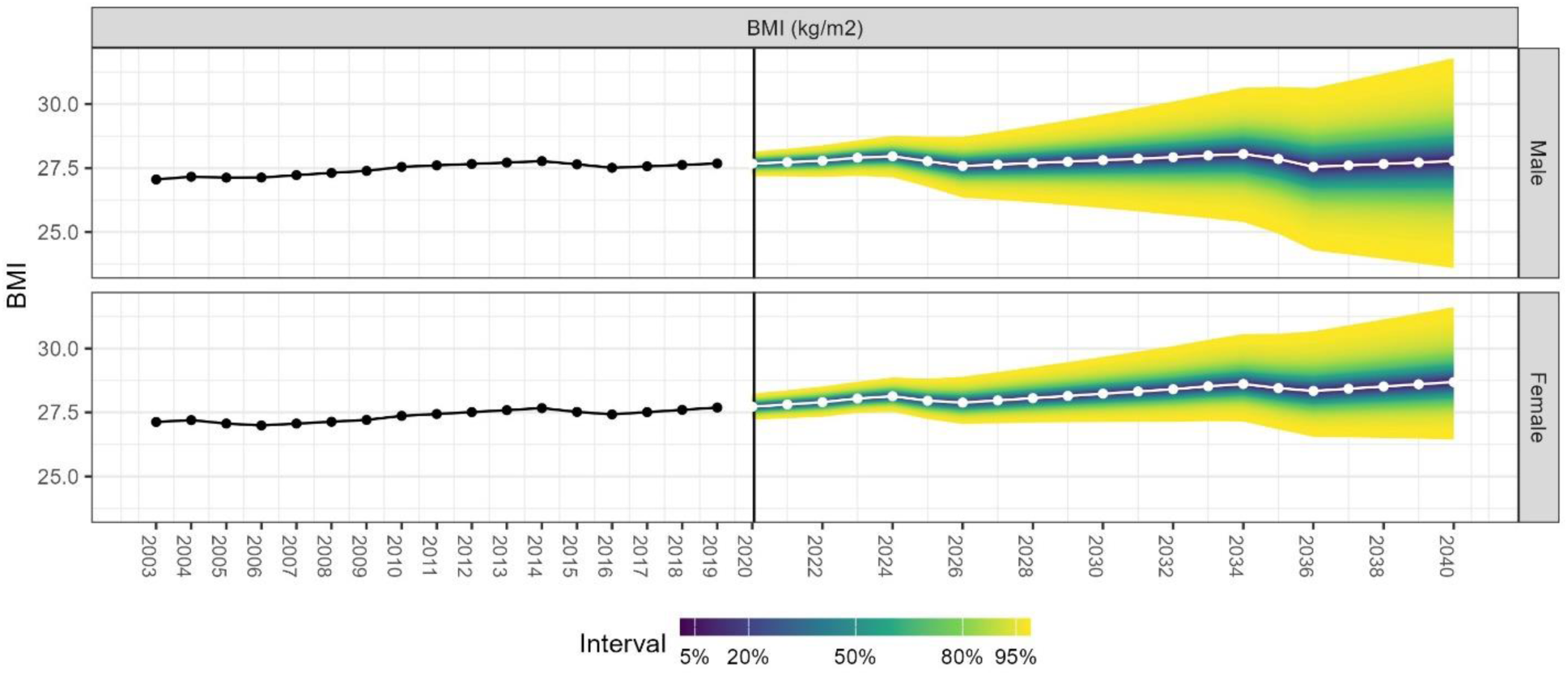
Sensitivity analysis with APC model assuming normal distribution of BMI in the population, by sex. BMI denotes ‘Body mass index’.

## Notes

### Competing Interest Statement

The authors have declared no competing interest.

### Author Declarations

An internal ethical review was carried out by Public Health Scotland which concluded that the study complied with the recognised ethical standards set out by the NHS Health Research Authority for all NHS health and social care research. Further approval was therefore not required from an independent NHS Research Ethics Committee.

